# Molecular evidence of coinfection with acute respiratory viruses and high prevalence of SARS-CoV-2 among patients presenting flu-like illness in Bukavu city, Democratic Republic of Congo

**DOI:** 10.1101/2021.10.16.21265044

**Authors:** Patrick Bisimwa Ntagereka, Rodrigue Ayagirwe Basengere, Tshass Chasinga Baharanyi, Théophile Mitima Kashosi, Jean-Paul Chikwanine Buhendwa, Parvine Basimane Bisimwa, Aline Byabene Kusinza, Yannick Mugumaarhahama, Dieudonné Wasso Shukuru, Simon Baenyi Patrick, Ronald Tonui, Ahadi Bwihangane Birindwa, Denis Mukwege

**Author notes:** **Corresponding author:** Patrick Bisimwa Ntagereka, Université Evangélique en Afrique, Molecular Biology Laboratory, Bukavu Democratic Republic of Congo, PO Box 3323.

## Abstract

The coronavirus 2019 (COVID-19) is caused by the severe acute respiratory syndrome coronavirus 2 (SARS-CoV-2) with clinical manifestation cases are almost similar to those of common respiratory viral infections. This study determined the prevalence of SARS-CoV-2 and other acute respiratory viruses among patients with flu-like symptoms in Bukavu city Democratic republic of Congo. We screened 1352 individuals with flu-like illnesses seeking treatment in 10 health facilities. Nasopharyngeal swabs specimens were collected to detect SARS-CoV-2 using real-time reverse transcription-polymerase chain reaction (RT-PCR) and 10 common respiratory viruses were detected by multiplex reverse transcription polymerase chain reaction assay. Overall, 13.9% (188/1352) patients were confirmed positive for SARS-CoV-2. Influenza A 5.6% (56/1352), and Influenza B 0.9% (12/1352) were the most common respiratory viruses detected. Overall more than two cases of the other acute respiratory viruses were detected. Frequently observed symptoms associated with SARS-CoV-2 positivity were shivering (47.8%; OR= 1.8; CI: 0.88-1.35), cough (89.6%; OR=6.5, CI: 2.16-28.2), myalgia and dizziness (59.7%; OR=2.7; CI: 1.36-5.85). Moreover, coinfection was observed in 12 (11.5%) specimens. SARS-CoV-2, and Influenza A were the most co-occurring infections, accounting for 33.3% of all positive cases. This study demonstrates cases of COVID-19 infections co-occurring with other acute respiratory infections in Bukavu city during the ongoing outbreak of COVID-19. These data emphasize the need for routine testing of multiple viral pathogens for better prevention and treatment plans.

## 1. INTRODUCTION

The world is facing a pandemic caused by the coronavirus disease 2019 (COVID-19), with over 136 million confirmed cases, including approximately 2,941,128 deaths globally as of Apr 13 2021^1^. COVID-19 is caused by severe acute respiratory syndrome coronavirus 2 (SARS-CoV-2), a novel coronavirus, that emerged in Wuhan, China, in December 2019^2^. SARS-CoV-2 infection is generally asymptomatic^3^, although in most confirmed cases, it may manifest itself with non-typical symptoms, including fever, cough, sore throat^3,4,5^, which are known to be present in common cold. Additionally, anosmia and ageusia may be observed^6,7^.

The Democratic Republic of the Congo (DRC), with an estimated population of 86 million, reported the first COVID-19 case on March 10, 2020, in Kinshasa city. While in Bukavu, a city whose population is estimated above 1.13 million, registered the first case of COVID-19 from arriving travelers in March 2020. Due to delay in quarantine measures and the failure in applying ministry of health guidelines for containment of the pandemic, DRC had recorded 55774 confirmed cases and 1061 deaths as of 4^th^ September 2021 in all 26 provinces, including South Kivu.

South Kivu is characterized by a high population density, poor health infrastructure and limited access to COVID-19 diagnostic tests. This places South Kivu among the high-risk provinces in DRC. Previous studies have reported high seroprevalence of SARS-Cov-2 among health workers and travelers in Bukavu city, suggesting a high circulation of SARS-CoV-2 within this city^8,9^. Diagnostic facilities for viral infections in DRC’s provinces, and in particular South Kivu are scarce. This prevents systematic screening of individuals who are asymptomatic or present mild symptoms of COVID-19 (https://ourworldindata.org/coronavirus-testing). This challenge further hampers strategies in the evaluation of the spread of SARS-CoV-2 within the population. Currently, COVID-19 ‘gold’ test, which entails reverse transcription-polymerase chain reaction (RT-PCR) as recommended by World Health Organization^3,10^, focuses on symptomatic cases, and hence, accurate estimate of the incidence cannot be obtained.

The research on the clinical characteristics of COVID-19 in most African countries has shown that fever and cough are the most common symptoms associated with COVID-19 diagnosis. Aside from these signs, fatigue, difficulty in breathing, sore throat, headache, and other atypical symptoms have been reported.

The glaring similarity between these clinical signs with those of other respiratory viral infections^11^ reduces the diagnostic efficiency and treatment of COVID-19 cases. This may lead to regular epidemics with pneumonia and bronchitis cases, severe respiratory failure, and even death^12^.

As the COVID-19 cases continue to rise, several cases of flu-like illness have been reported in Bukavu city by the Provincial Ministry of Health from December 2020 to April 2021.

Therefore, identifying SARS-CoV-2 and pathogens responsible for respiratory infections is key in the prevention of COVID-19 spread. This could improve patient management and reduce patient isolation duration, particularly for those infected with other common respiratory viruses.

Thus, this study investigated the prevalence of SARS-CoV-2, Influenza A and B, and other acute respiratory viruses among local patients with flu-like symptoms. These include but were not limited to patients presenting a sudden onset of a fever of (>38 °C) and a cough or sore throat who were seeking treatment at different health structures in Bukavu city.

## 2. METHODS

### 2.1 Study area

This study was carried out in Bukavu, the capital city of South Kivu province in eastern DRC. The city is lying at the extreme south-western edge of Lake Kivu and separated from Rwanda by the outlet of the Ruzizi River. As of 2020, Bukavu had an estimated urban population of 1,133,371. (https://populationstat.com/democratic-republic-of-the-congo/bukavu). Moreover, the city is characterized by a tropical wet and dry climate with 1498 m of altitude above sea level and an average rainfall of about 1224 mm annually. Patients were recruited from December 2020 to May 2021 in 10 health facilities such as BIOPHARM Health Centre, Panzi General Reference Hospital, Saint Vincent polyclinic, MUHUNGU Health Centre, DIOCESAIN Centre, Saint Luc polyclinic, 5^th^ CELPA Health Centre, SOS Medical Centre, Nyantende Hospital, and CHAI Health Centre.

### 2.2 Study design, participants, and sample collection

A cross-sectional study was adopted. Collection of swab samples to confirm COVID-19 disease among patients with flu-like symptoms defined as an outpatient with a sudden onset of a fever >38 °C, a cough or sore throat seeking treatment at the different health facilities. Only individuals with flu-like disease symptoms, aged ten years and above, and able to give consent were considered for sample collection the interview. Personal data such as age, sex, occupation, and residence were recorded prior to collecting oropharyngeal swab samples. Collected samples were put in a 2ml tube containing 2 milliliters of Sample Storage Reagent (Ansure, Biotechnology) composed of 0.9% of normal saline raisin, and stored in -40°C before SARS-CoV-2 Reverse Transcriptase Polymerase Chain Reaction amplification.

A total of 1352 Oropharyngeal samples were collected and shipped daily in ice to the Molecular Biology laboratory of the Université Evangélique en Afrique (UEA). The data collection followed ethical guidelines applied for human biological data collection and processing during the study.

### 2.3 Molecular detection of SARS-CoV-2

Viral RNA was extracted from swabs samples within 1–24 hours following sample collection using innuPREP Virus TS RNA kit (Analytik Jena, Berlin, Germany) according to the manufacturer’s instructions. The real-time reverse transcriptase-polymerase chain reaction (RT-PCR) tests for SARS-CoV-2 were performed using TIB MOLBIOL Sarbeco E-gene Plus EAV PCR Kit (Olfert Landt, Berlin Germany). Briefly, this entailed a 20 μL reaction mix comprising 10 μL of 2x Reverse transcriptase master Mix, 0.5 μL of extraction control EAV, 0.5 μL of PSR reagent (SARS-CoV-2 probes and primers mix targeting the viral E gene), 5 μL of the extracted RNA and the volume was adjusted to 20 μL with nuclease free water.

The cDNA synthesis step was conducted by incubating the mixture at 55°C for 5 minutes. RT-PCR reactions with an initial denaturation step at 95°C for 5 min, followed by cycling steps of 95°C for 5 seconds, primer annealing at 60°C for 15 seconds, and an extension step of 72°C for 15 seconds. All the RT-PCR reactions were carried out in LightCycler^®^ 96 Instrument (Roche, Life Science). A sample was considered positive if the threshold (Ct value) is *<*35 while any sample with threshold (Ct value) >35 was considered as negative.

### 2.4 Detection of common acute respiratory viruses by Multiplex RT-PCR assays

Each RNA sample was subjected to multiplex RT-PCR amplification using the ARVI Screen Real-TM Multiplex Detection kit (Sacace Biotechnology, Italy) on a Lightcycler 96 detection system (Roche, Germany). The panel was specifically developed to identify ten viruses including human respiratory syncytial virus (hRSV); human metapneumovirus (hMpv); human parainfluenza virus-1-4 (hPiv-1-4), OC43, E229, NL63 and HKUI human coronavirus (hCov); human rhinovirus (hRv); human B, C, and E adenovirus (hAdv), human bocavirus (hBov) as well as Influenza A and B within 1hour. The assay comprised reverse transcription and multiplex RT-PCR Amplification.

The cDNA synthesis was performed in a 40 μL mix containing 5 μl RT-G-mix-1, 6μl of Reverse transcriptase (M-MLV), 9 μL of RT-Mix, and 20 μL of RNA as a template. The cDNA libraries were constructed by incubating the reaction mix at 37°C for 30 minutes in PeQlab thermocycler, (peqSTAR, VWR). The cDNA samples were diluted in half with T.E. buffer for immediate use in multiplex RT-PCR amplification or storage at -20°C for future analysis.

In addition, the multiplex Real-Time amplification setup was carried. Each mix contained primers directed against regions of acute respiratory viral infection (ARVI). For each mix, three controls were also included, such as negative, positive and internal control.

Real-time RT-PCR assays were performed in a 25 μL reaction comprising 10 μL of PCR-mix 1 containing specific primers for each virus, 5 μL of PCR-Mix-FRT and 10 μL of template cDNA. The cycling parameters entailed 95 °C initial denaturation for 15 min followed by 40 cycles of 95 °C denaturation for 10s, 54°C annealing for 25s and extension at 72 °C for 25s. We set our threshold value for positive cases of ARVI pathogens at any value less than 35.

The negative cases of an ARVI were assessed when the threshold value (Ct) of the tested sample is absent (not determined) or if the Ct value exceeds 35.

### 2.5 Data analysis

Collected data were encoded in Microsoft^®^ Excel^®^ data management tool combining the results of SARS-CoV-2 RT-PCR and other respiratory virus assays and data obtained from the questionnaire. Prior to statistical analyses, all personal identification data were eliminated. Descriptive statistics were calculated for all of the variables, including the RT-PCR prevalence results. We assessed the associations between SARS-CoV-2 and other respiratory viruses RT-PCR results (positive or negative) with age, sex, symptom, exposure and morbidity using the Chi-Squared test. The odds of being a positive case based on RT-PCR results were then modeled as a function of the dichotomous risk factors measures, using logistic regression models. All variables with p-values of ≤0.05 were considered statistically significant. The R. Console statistic software (version 4.0.0) was used to perform the analysis.

## 3. RESULTS

### 3.1 SARS-CoV-2 and common acute respiratory infection prevalence and associated Symptoms in patients with flu-like symptoms from nine health structures

In total, 1352 individuals presenting flu-like illness were examined, all of whom resided in the Bukavu city. Among these were 696 males (51.4%) and 656 females (48.5%). The majority (89.6%) being adults. A high proportion (63.3%) were those recorded in BIOPHARM medical center. The SARS-CoV-2 was detected in 188 representing a positivity rate of 13.9%. The prevalence of SARS-CoV-2 was higher in females (14.6 %) compared to males (13.3%). The age range of the SARS-CoV-2-positive patients was 176(14.5%) out 1212 adults and 12(8.6%) in young (less than 18 years).out of 140.

Influenza A and B viruses were diagnosed respectively in 76 (5.6%) and in 12 (0.9%) samples. Multiplex RT-PCR test detected Human parainfluenza 1-4 (hPiV 1-4) in 10 (0.7%) patients while human Metapneumonia virus (hMpv) was present in 5 (0.4%) patients. Additionally, human Coronavirus (hCoV), human Rhinovirus (hRV) and human Adenovirus (hAdV) were identified in 3 (0.2%) samples for each while only 1(0.07%) patient was identified to be infected with Human Bocovirus (hBoV).

Of all health facilities, Panzi General Reference (Panzi HGRP) recorded the highest SARS-CoV-2, 12 (23.1%), and influenza B 4 (7.7%) cases (Table 1).

**TABLE 1.** Frequency of SARS-CoV-2 and other respiratory viruses detected according to sex, age and health facility

| Variables | Number<br>sampled | SARS-CoV-<br>2 + | INFL. A + | INFL. B + | hMpv | hPiV 1-4 | hCoV | hRV | hAdV | hBoV |
| --- | --- | --- | --- | --- | --- | --- | --- | --- | --- | --- |
| **Gender** |  |  |  |  |  |  |  |  |  |  |
| F | 696 | 101(14.6%) | 40(5.8 %) | 8(1.2 %) | 3(0.4 %) | 7(1.0 %) | 1(0.1 %) | 2(0.3%) | 1(0.1%) | 1(0.1%) |
| M | 656 | 87(13.3%) | 36(5.5%) | 4(0.6 %) | 2(0.3 %) | 3(0.5 %) | 2(0.3 %) | 1(0.2%) | 2(0.3 %) | 0(0 %) |
| **Age range** |  |  |  |  |  |  |  |  |  |  |
| Adult | 1212 | 176(14.5%) | 60(5.0%) | 12(1.0 %) | 4(0.3 %) | 8(0.7 %) | 3(0.2 %) | 3(0.2) | 3(0.2 %) | 1(0.1 %) |
| Young | 140 | 12(8.6%) | 12(8.6%) | 0(0 %) | 1(0.7 %) | 2(1.4 %) | 0(0%) | 0 (0%) | 0(0%) | 0(0%) |
| **Health structures** |  |  |  |  |  |  |  |  |  |  |
| Biopharm HC | 856 | 116(13.6%) | 48(5.6%) | 4(0.5 %) | 1(0.1 %) | 4(0.5 %) | 0(0 %) | 0(0%) | 0(0%) | 1 (0.1%) |
| 5<sup>th</sup> CELPA HC | 72 | 8(11.1%) | 4(5.6 %) | 0(0 %) | 1 (1.4%) | 2(2.8%) | 1(1.4 %) | 1(1.4%) | 2(2.8%) | 0(0%) |
| Panzi GRH | 52 | 12(23.1%) | 0(0%) | 4(7.7%) | 2(3.8%) | 0(0%) | 2 (3.8 %) | 0 (0%) | 0(0%) | 0(0%) |
| CHAI HC | 64 | 4(6.3%) | 4 (6.3 %) | 0(0 %) | 0 (0) | 0 (0 %) | 0 (0 %) | 0 (0 %) | 0(0 %) | 0(0 %) |
| SOS HC | 88 | 16(18.2%) | 4(4.5%) | 0 (0%) | 0 (0 %) | 0 (0 %) | 0 (0 %) | 0 (0 %) | 0(0%) | 0(0%) |
| Nyantede Hospital | 76 | 12(15.8%) | 4(5.3%) | 0(0 %) | 0 (0 %) | 0(0 %) | 0 (0 %) | 1(1.3 %) | 0(0%) | 0(0%) |
| Muhungu dios.HC | 20 | 4(20.0 %) | 0(0%) | 0(0%) | 0(0%) | 0 (0%) | 0(0%) | 0(0%) | 0(0%) | 0(0%) |
| St Luc HC | 24 | 8(33.3 %) | 4(16.7 %) | 0(0 %) | 1 (4.2%) | 2(8.3%) | 0(0%) | 1(4.2%) | 1(4.2%) | 0(0%) |
| Muhungu HC | 28 | 8(2.9 %) | 4(1.4 %) | 4(1.4%) | 0(0%) | 2 (0.7%) | 0(0%) | 0(0%) | 0(0%) | 0(0%) |
| **Total N (%)** | **1352** | **188 (13.9)** | **76 (5.6)** | **12 (0.9)** | **5(0.4)** | **10(0.7)** | **3(0.2)** | **3(0.2)** | **3(0.2)** | **1(0.07)** |

### 3.2 Profiles of SARS-CoV-2 and other respiratory infection positive patients and associated clinical symptoms

The most frequent symptom in COVID-19 was coughing (89.6%), followed by headaches 1049 (77.6%), asthenia 909 (67.2%), high fever 706 (52.2%), as well as myalgia (59.7%). Hypertension 122(9%), obesity 81(6%), and diabetes 81(6%) were the most frequently reported comorbidities in COVID-19 positive individuals. More than half of influenza A-positive patients experienced fever, cough, and headache and were asthenia, hypertensive and diabetics. However, all influenza B infected patients experienced headaches (100%) while more than half presented fever, coldness, cough, breathing difficulties, asthenia, myalgia, anorexia, and sputum **(Supplementary materials Table 1)**. The statistical analysis revealed statistically significant association between COVID-19 with some symptoms such as shivering (47%.8%; OR= 1.8; CI: 0.88-1.35; p=0.024), cough (89.6%; OR=6.5, CI=2.16-28.2; p=0.003), myalgia and dizziness (59.7%; OR=2.7; CI: 1.36-5.85, p=0.006) (Table 2).

**TABLE 2:** Association of flu-like symptoms with SARS-CoV-2 positivity among flu-like patients seeking treatment in different health facilities

| Variables | Negative covid19<br>N= 1164(%) | Positive<br>Covid19<br>N= 188(%) | OR (CI) | p value |
| --- | --- | --- | --- | --- |
| **Symptoms (%)** |  |  |  |  |
| **Fever** | 512 (44.1%) | 98 (52.2%) | 1.1 (0.560-2.282) | 0.734 |
| **Coldness** | 531 (45.6%) | 98 (52.2%) | 0.9 (0.456- 1.856) | 0.816 |
| **Shivering** | 371 (31.9%) | 88 (47.8%) | 1.8 (0.88 - 1.35) | 0.0246** |
| **Sweating** | 268(23.0%) | 56 (29.9%) | 1.3 (0.603- 3.69) | 0.452 |
| **Cough** | 764(65.6%) | 168 (89.6%) | 6.5 (2.161- 28.207) | 0.003*** |
| **Breathing difficulties** | 271(23.3%) | 45 (23.9%) | 0.7 (0.321-1.814) | 0.590 |
| **Essoufflés** | 233(20.0%) | 17 (9.0%) | 0.3(0.107-1.094) | 0.0992 |
| **Headaches** | 793(68.1%) | 146 (77.6%) | 1.6 (0.738-3.821) | 0.242 |
| **Asthenia** | 560(56.7%) | 126 (67.2%) | 1.6(0.804-3.487) | 3.487 |
| **Myalgia** | 478(41.1%) | 112 (59.7%) | 2.7(1.363-5.859) | 0.006*** |
| **Anorexia** | 314(27.0%) | 73 (38.8%) | 1.5 (0.743-3.283) | 0.232 |
| **Sputum** | 194(16.7%) | 42 (22.4%) | 0.8(0.360-1.926) | 0.716 |
| **Dyspnea** | 164(14.1%) | 19 (10.4%) | 0.6 (0.236-1.772) | 0.461 |
| **Expositions (%)** |  |  |  |  |
| **Stay in high risk areas** | 116 (10.0) | 33 (17.9) | 1.09 (0.31- 2.81) | 0.85 |
| **Close contact** | 48 (4.1) | 11 (6.0) | 2.40 (0.43-13.45) | 0.29 |
| **Work in hospitals** | 81 (7.0) | 17 (9.0) | 1.78 (0.34-8.43) | 0.46 |
| **Comorbidities (%)** |  |  |  |  |
| **HIV infection** | 8 (0.7) | 0 (0.0) | - | - |
| **Arterial hypertension** | 112 (9.6) | 17 (9.0) | 0.82 (0.21-2.57) | 0.75 |
| **Obesity** | 30 (2.6) | 11 (6.0) | 0.76 (0.03-6.16) | 0.81 |
| **Diabetes** | 69(5.9) | 11 (6.0) | 1.17 (0.29-3.93) | 0.80 |
| **Chronic respiratory dis.** | 26 (2.2) | 3 (1.5) | 0.56 (0.02-3.97) | 0.61 |
| **Cardio pathology** | 8 (0.7) | 0 (0.0) | - | - |
| **Chronic kidney dis.** | 13 (1.1) | 0 (0.0) | - | - |
| **Nero pathology** | 5 (0.4) | 1.5 | 0.00 (0.00-0.001) | 0.98 |

\*\*\*: highly significant; \*\*: Very significant; \*: Significant

Comparative analysis between SARS-CoV-2 positive and negative individuals showed that patients who have been in contact with infected persons presented a higher risk of being infected by SARS-CoV-2 were the one with close contact (OR=2.40; CI: 0.43-13.45), followed workers in hospitals (OR=1.78; CI: 0.34-8.43) and patient with diabetes (OR=1.17; CI: 0.29-3.93) than negative individuals. However, none of these factors were statistically significant. Additionally, no statistical difference was found between gender and commodities variables when compared SARS-CoV-2 and negative patients (p<0.05) (Table 2).

### 3.3 Logistic regression analysis for factors associated with Influenza A and B positivity

The logistic regression analysis revealed that 53.3% (OR=1.51; IC=0.46-4.98) of Influenza A negative patients were inversely correlated with cold compared with 16.7% in influenza A positive patients. However, there were no significant differences in other factors for influenza A and B infection rates (*p > 0*.05) (Table 3).

**TABLE 3:**
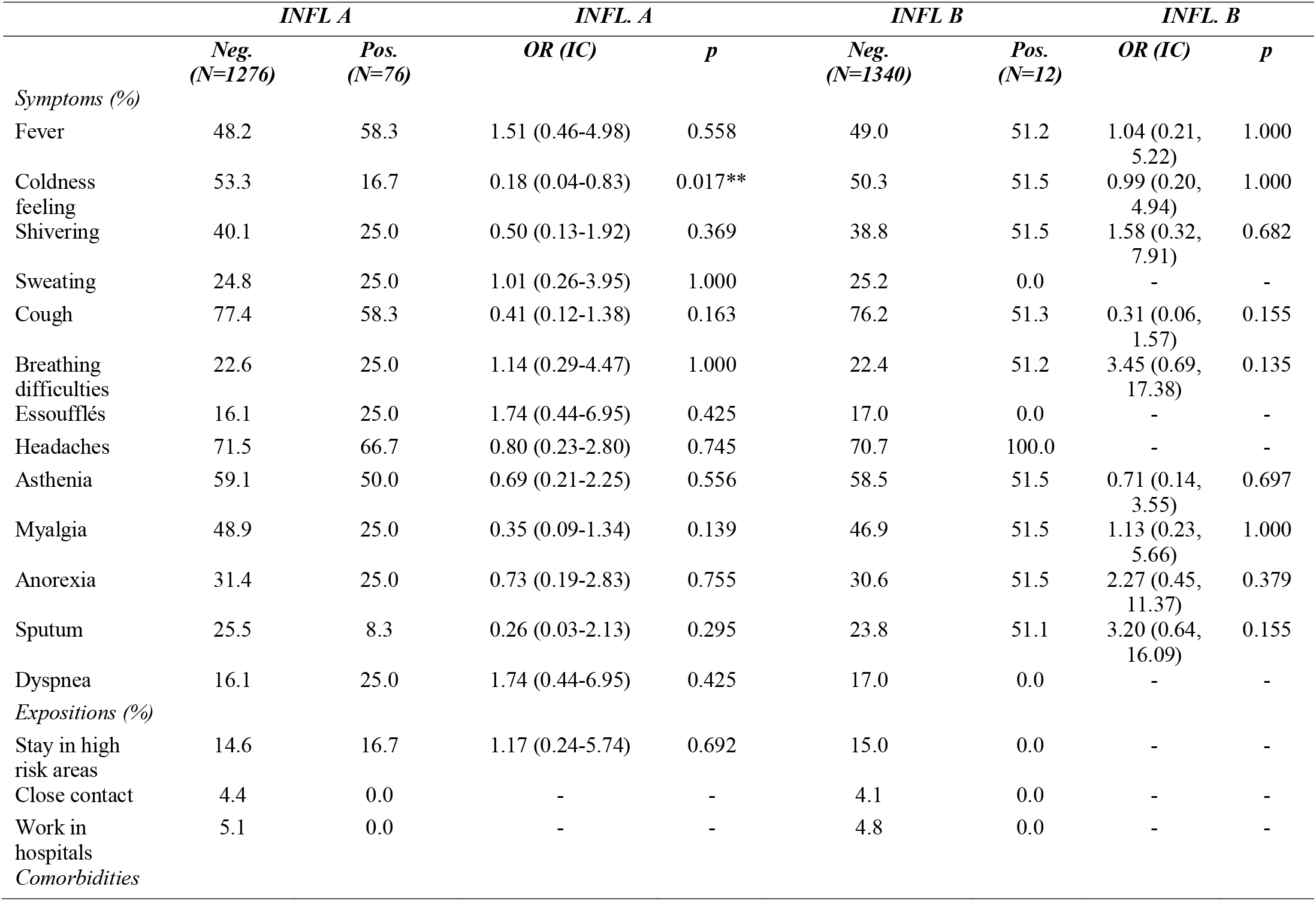
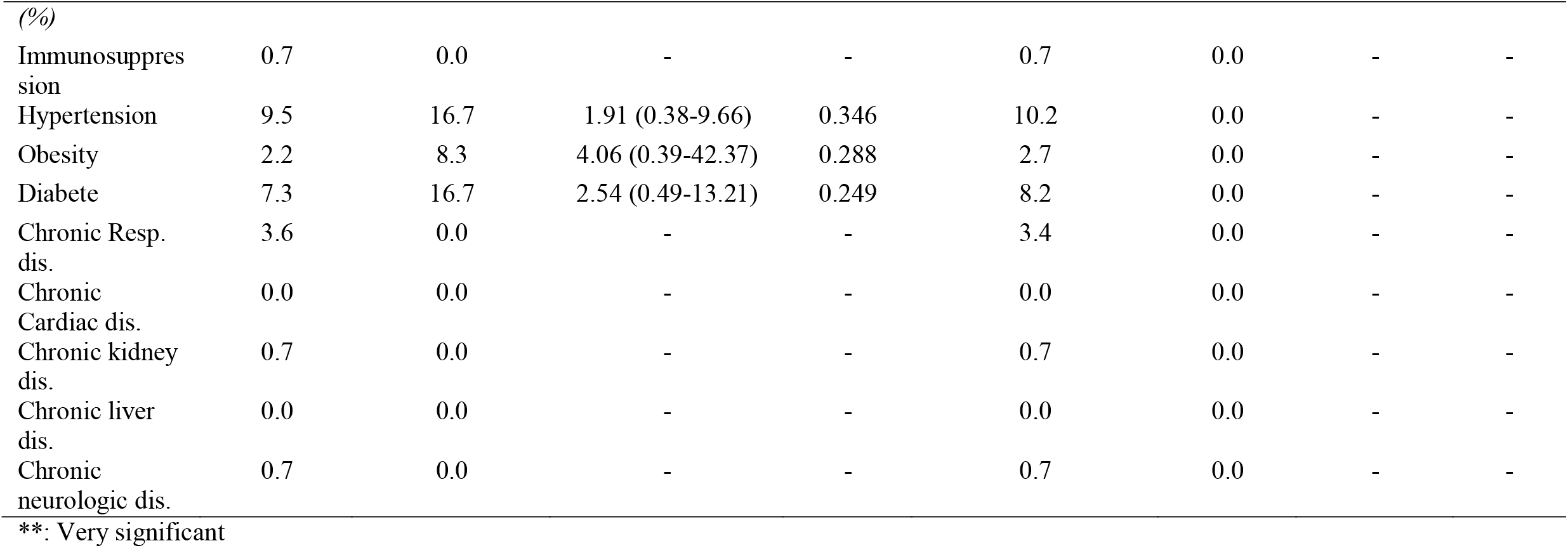
Logistic regression analysis regarding demographical gender, exposition and comorbidity between positive Influenza A and B patients

### 3.4 Coinfection patterns of COVID-19 and other acute respiratory viruses

During this investigation, it was found six coinfection patterns. Out of the 113 cases of infected acute respiratory patients, 12 (11.5%) cases were co-infected with different acute respiratory viruses. A high frequency of coinfections pathogens involved SARS-CoV-2 and Influenza A, accounting for 33.3% of all coinfection cases. Additionally, 3 cases (25%) of hMPiV positive cases were co-infected with hCoV followed by 2 cases (16.6%) of SARS-CoV-2 co-infected with Influenza B (Table 4).

**TABLE 4:** Co-infection of SARS-CoV-2 with others respiratory viruses in Flu-like symptomatic patients

| *Number* | *Virus types* | *Co-infectious viruses* | *Cases* |
| --- | --- | --- | --- |
| 1 | hMPV+hPiV 1-4 | 2 | 1 |
| 2 | hMPV+hCoV | 2 | 3 |
| 3 | HBoV+Inf. A | 2 | 1 |
| 4 | SARS-CoV-2+Inf. B | 2 | 2 |
| 5 | SARS-CoV-2+Inf. A | 2 | 4 |
| 6 | SARS-CoV-2+hMPiV | 2 | 1 |

## 4. DISCUSSION

COVID-19 first reports in DRC emerged towards the beginning of March 2020 and there has been an upsurge of the cases ever since then nationally. SARS-COV-2 infections in South Kivu province soared from April to December 2020. This is the period when flu-like illnesses peaks. It was therefore vital to investigate the presence of SARS-CoV-2 among local patients with flu-like illnesses seeking treatment in various health facilities. This may help us to better control the epidemic outbreak and correctly handle other acute respiratory infections. Participants in this study included 696 males (51.4%) and 656 females (48.5%). The majority were adults 1212 (89.6%).

In this study, a total of 188 participants were confirmed to be COVID-19 positives with positivity rate of 13.9%. The infection rate found from this study is higher than the one found in a previous study conducted in China where only 0.3% of patients with influenza-like illnesses were found to be COVID-19 positive^13^. Similarly, these results are not consistent with the ones reported in South Kivu, DRC, in which SARS-CoV-2 antibodies were detected in 148 (41.2%) health workers from the Panzi General Reference Hospital (HGRP)^9^. This difference could probably be explained by the type of test and study population. The previous study from HGRP was detecting only antibodies (exposure) and focusing only on asymptomatic health workers while in this study, we examined individuals presenting flu-like illness seeking treatment in different health facilities.

In addition, 8.3% of the flu-like illness patients were tested positive for other respiratory viruses. These results emphasize the importance of simultaneous diagnosis of other acute respiratory viral pathogens, particularly in a mild epidemic area and during the current COVID-19 pandemic. Among the acute respiratory viral pathogens, Influenza A and influenza B viruses were the most detected respectively in 76 (5.6%) and 12 (0.9%) patients with flu-like symptoms and COVID-19 negative. This infection rate is relatively lower than those reported in Shanghai in which influenza A and B were detected in 40.4% and 14.3% of patient respectively^14^. In Chongqing, 65.7% cases of Influenza A and 34.1% of Influenza B were reported in individuals with flu-like symptoms^15^. This differences could be explained by the difference in targeted population, the previous was focused mostly while in the last study the majority were children and young people. As the considered area face different agro-ecological conditions, this may lead to different rate of infection. However, findings from this study are in agreement with a recent study revealing a higher incidence of influenza infections in children and adolescents compared to old individuals^16^.

Furthermore others respiratory pathogens were detected including hPiV 1-4, hPiV 1-4 10(0.7%), hMPiV 5(0.4) and the other virus with 3 cases (0.2%) each. Similar results were obtained in different studies in Costa Rican and China^17^. In all the cases, adult persons tended to be mostly affected compared to young people although no statistical differences were found.

A weak association was noted in patients infected with respiratory viruses and clinical signs such as cough, headaches, asthenia, coldness feeling, fever, shivering, and myalgia. This result is in agreement with the finding from Monto et al.^18^ that revealed cough associated with fever to be the most frequent symptoms with influenza infection. However, our findings are inconsistent with a previous study focused on clinical features of COVID-19 and Influenza, which demonstrated that symptoms like anosmia, dysgeusia, diarrhea, frontal headache and bilateral cracklings sounds were more frequent in COVID-19 while sputum production, dyspnea, sore throat, conjunctivitis, hyperthermia, tearing, vomiting and rhonchi sounds were more frequent in influenza infection^19^. A subsequent study with a larger sample and followe up of patients would allow to establish clinical signs of similarities and differences between infection with COVID-19 and infection with other respiratory viruses. This can help practically in the areas under equipped for the biological diagnosis.

In this study, gender and comorbidities did not significantly influence the COVID-19 rate. This is similar with previous studies showed that gender was not associated with the SARS-CoV-2 infection rate^20, 21^.

The risk of being infected by SARS-CoV-2 increased in patients who have been in contact with diseased persons (OR=2.4). This is in accordance with the fact that in resource-limited areas, including South Kivu province, quarantine of infected individuals often occurred at home, enabling onwards transmission within households.

People of all ages are susceptible to SARS-CoV-2. However, previous findings have confirmed that adults, specifically older men are more likely to be affected and subsequently suffer from severe pneumonia, pulmonary oedema, acute respiratory distress syndrome, multiple organ failure, and death^5, 22,23^. Although it appeared that all ages of the population are susceptible to SARS-CoV-2 infection, but the median age of infection was in adult persons (around 50 years). This is in agreement with previous research revealing the median age of infection to be around 50 years^20^, and considering the range of African population.

In general, there are no striking differences in the clinical behavior and severity between the different Influenza A and B viruses affecting patients. The only significant clinical symptom with negative relationship to Influenza A virus was feeling cold (OR= 0.18, p=0.017) where the majority of influenza A negative individuals were feeling cold. Our data do not show differences in clinical manifestation compared with other previous descriptions in the literature. However, three clinical symptoms were relevant for Influenza A, including fever, cough, headache, feeling cold. Similar results were reported in a previous study in Costa Rica^16^. Among the 12 patients who were Influenza A type positive in our study, all (100%) had a headache, and more than half presented the remaining symptoms such as Fever, Coldness, Shivering, Sweating, Cough, Breathing difficulties, essoufflés, Asthenia, myalgia, anorexia, sputum, and Dyspnea. No respiratory and cardiac failure was observed to be associated with Influenza infection. This result is in contrast with previous studies that have reported 16% of cases of seasonal Influenza occurring in clinical pulmonary patients^24^ and 25.2% in cardiac failure^25^.

Moreover, 12 (11.5%) cases co-infected with other acute respiratory viruses were reported. The highest coinfections pathogens were SARS-CoV-2 and Influenza A, accounting for 33.3% of all coinfection cases. The high coinfection rate between SARS-CoV-2 and Influenza virus can probably be explained by restrictions in applying barrier measures such as wearing masks, hand washing, social distance, and hand-sanitizing of infected person. This could further be due to the seasonality nature of the influenza virus which peaks between December and April, corresponding to the winter period. This observation is supported by a previous study conducted by Leung et al.^26^. However, our finding is in contrast with previous studies^27,28^ whose findings show a decrease in seasonal influenza cases during the Covid-19 pandemic. In this regard, it would be ideal to have the follow-up of patients co-infected with COVID-19 and other respiratory viruses and to assess their evolution over time; this may also requires another extended study for clinical follow-up of cases.

In addition, in this study, cases of hMPiV co-infected with hCoV were reported. These findings are in accordance with previous studies suggesting the possible coinfections of multiple respiratory viruses^29, 30, 31,32^.

This study demonstrates cases of COVID-19 infections co-occurring with other acute respiratory infections in DR Congo. These data on the coinfection of SARS-CoV-2 with respiratory viral pathogens emphasize the need for routine testing of multiple viral pathogens in mild epidemic areas during the current COVID-19 epidemic. Further study is need to evaluate clinical patient with these co-infection and their evolution. Additionally, whether simultaneous viral infection in SARS-CoV-2 patients can potentially drive viral interference or impact disease outcome need further investigation.

## 5. CONCLUSION

This study analysed the epidemiological patterns of SARS-CoV-2 and different respiratory viruses, and found a high infection rate with COVID-19 and cases of positive common acute respiratory infections in patients presenting flu-like symptoms. Although the current pandemic constitutes a serious public health concern, SARS-CoV-2 may not be considered to be the only pathogen responsible for respiratory tract infection in South Kivu eastern of DRC. Therefore, testing for respiratory viruses should be performed in all patients with respiratory symptoms for effective surveillance of the transmission patterns in the COVID-19 affected areas for optimal treatment to inhibit the rapid spread of the virus

## Data Availability

All data produced in the present study are available upon reasonable request to the authors

## ETHICS STATEMENT

The study protocol was approved by the Interdisciplinary Center of Ethical Research (CIRE) of the Université Evangélique en Afrique (UEA), Ref: CNES 001/DPSK/115PP/2021. Informed consent was obtained from all participants after being signed and the confidentiality of the data was ensured.

## AUTHOR CONTRIBUTIONS

Patrick Bisimwa Ntagereka completed study design, project administration, data collection, drafted the manuscript.

Rodrigue Ayagirwe Basengere completed investigation, data curation, review and the editing the manuscript.

Tshass Chasinga Baharanyi completed investigation, and review of the manuscript.

Théophile Mitima Kashosi completed data curation, review and editing of the manuscript.

Jean-Paul Chikwanine Buhendwa completed investigation, data collection review and editing.

Parvine Basimane Bisimwa completed conceptualization, writing, and original draft preparation.

Aline Byabene Kusinza, investigation, data collection review and editing of the manuscript Yannick Mugumaarhahama completed formal analysis, visualization, writing, and original draft preparation.

Dieudonne Wasso Shukuru completed conceptualization, data analysis, writing, review, and editing.

Simon Baenyi Patrick completedformal analysis, data curation, writing, and original draft preparation.

Ronald Tonui, completed formal analysis, visualization, writing, and original draft preparation.

Ahadi Bwihangane Birindwa completed investigation, data curation, review and editing of the manuscript.

Denis Mukwege, supervised the study, completed project administration, writing, review, and editing the manuscript. All authors approved the manuscript.

## ACKNOWLEDGMENTS

We Acknowledge the Université Evangélique en Afrique (UEA) through the Bioscience unit and German Institute for Medical Mission (DIFÄM) for the financial support. Our sincere appreciation goes further to the Provincial Ministry of Health and all the leaders of health structures for facilitating access to sample and data. We also thank you the lab technicians of the Molecular Biology laboratory of UEA for their technical support.

