## Supplementary material for "Molecular evidence of coinfection with acute respiratory viruses and high prevalence of SARS-CoV-2 among patients presenting flu-like illness in Bukavu city, Democratic Republic of Congo": Suplementary material table 1

**Supplementary material table 1:** Characteristics of the overall flu-like SARS-CoV-2 positive patients and others respiratory viruses

|  | ***SARS-CoV-2 N= 1352*** | | ***INFL A 1352*** | | ***INFL B 1352*** | | ***Others Resp. viruses 1352*** | |
| --- | --- | --- | --- | --- | --- | --- | --- | --- |
|  | ***Negative %*** | ***Positive %*** | ***Negative %*** | ***Positive %*** | ***Negative %*** | ***Positive %*** | ***Negative %*** | ***Positive %*** |
| ***GENDER (%)*** |  |  |  |  |  |  |  |  |
| F | 81.8 | 18.2 | 93.1 | 6.9 | 97.3 | 2.7 | 96.15 | 3.85 |
| M | 80.1 | 19.9 | 90.8 | 9.2 | 100.0 | 0.0 | 97.33 | 2.67 |
| AGE | 37.5±16.9 | 40.9±13.7 | 39.2±15.9 | 43.5±18.2 | 39.5±16.1 | 43.0±9.9 |  |  |
| ***SYMPTOMS (%)*** |  |  |  |  |  |  |  |  |
| FEVER | 44.1 | 52.2 | 48.2 | 58.3 | 49.0 | 51.2 | 97.92 | 2.07 |
| COLDNESS FEELING | 45.6 | 52.2 | 53.3 | 16.7 | 50.3 | 51.5 | 96.45 | 3.55 |
| SHIVERING | 31.9 | 47.8 | 40.1 | 25.0 | 38.8 | 51.5 | 97.34 | 2.66 |
| SWEATING | 23.0 | 29.9 | 24.8 | 25.0 | 25.2 | 0.0 | 94.97 | 5.03 |
| **COUGH** | 65.6 | 89.6 | 77.4 | 58.3 | 76.2 | 51.3 | 97.34 | 2.66 |
| BREATHING DIFFICULT. | 23.3 | 23.9 | 22.6 | 25.0 | 22.4 | 51.2 | 98.52 | 1.48 |
| ESSOUFFLES | 20.0 | 9.0 | 16.1 | 25.0 | 17.0 | 0.0 | 96.74 | 3.26 |
| HEADACHES | 68.1 | 77.6 | 71.5 | 66.7 | 70.7 | 100.0 | 96.15 | 3.85 |
| ASTHENIA | 56.7 | 67.2 | 59.1 | 50.0 | 58.5 | 51.5 | 97.04 | 2.96 |
| **MYALGIA** | 41.1 | 59.7 | 48.9 | 25.0 | 46.9 | 51.5 | 97.34 | 2.66 |
| ANOREXIA | 27.0 | 38.8 | 31.4 | 25.0 | 30.6 | 51.5 | 96.74 | 3.26 |
| SPUTUM | 16.7 | 22.4 | 25.5 | 8.3 | 23.8 | 51.1 | 97.93 | 2.07 |
| DYSPNEA | 14.1 | 10.4 | 16.1 | 25.0 | 17.0 | 0.0 | 97.93 | 2.07 |
| ***EXPOSITIONS (%)*** |  |  |  |  |  |  |  |  |
| STAY IN HIGH-RISK AREAS | 10.0 | 17.9 | 14.6 | 16.7 | 15.0 | 0.0 | 97.93 | 2.07 |
| CLOSE CONTACT | 4.1 | 6.0 | 4.4 | 0.0 | 4.1 | 0.0 | 99.70 | 0.30 |
| WORK IN HOSPITALS | 7.0 | 9.0 | 5.1 | 0.0 | 4.8 | 0.0 | 98.52 | 1.48 |
| ***COMORBIDITIES (%)*** |  |  |  |  |  |  |  |  |
| HIV infection | 0.7 | 0.0 | 0.7 | 0.0 | 0.7 | 0.0 | 100 | 0 |
| HYPERTENSION | 9.6 | 9.0 | 9.5 | 16.7 | 10.2 | 0.0 | 98.52 | 1.48 |
| OBESITY | 2.6 | 6.0 | 2.2 | 8.3 | 2.7 | 0.0 | 99.70 | 0.30 |
| DIABETES | 5.9 | 6.0 | 7.3 | 16.7 | 8.2 | 0.0 | 99.12 | 0.88 |
| Chronic Resp. dis | 2.2 | 1.5 | 3.6 | 0.0 | 3.4 | 0.0 | 99.11 | 0.89 |
| Cardio pathol. | 0.7 | 0.0 | 0.0 | 0.0 | 0.0 | 0.0 | 100 | 0 |
| Chronic kidney dis. | 1.1 | 0.0 | 0.7 | 0.0 | 0.7 | 0.0 | 100 | 0 |
| Neuro pathol. | 0.4 | 1.5 | 0.7 | 0.0 | 0.7 | 0.0 | 100 | 0 |
